# ‘Effectiveness of dual-mobility cups for preventing dislocation after primary total hip arthroplasty by a posterolateral approach and their cost-effectiveness compared to unipolar cups in elderly patients.’

**DOI:** 10.64898/2026.08.18.26360681

**Authors:** Okke Hoonhout, Esther Scheijbeler, Loes W A H van Beers, Dirk Jan F Moojen, Amanda D Klaassen, Nienke W Willigenburg, Rudolf W Poolman, REDEP collaborators

## Abstract

**Rationale:** Dislocation is the leading reason for early revision surgery. To address the problem of dislocation, the dual-mobility (DM) cup was developed in France in the 1970’s. This cup should provide more stability and biomechanically reduce the risk of dislocation. In the Netherlands, most DM cups are placed in specific patients, e.g. with cognitive impairment and for revisions due to recurrent dislocations. Despite the increased and, in some countries, broad use of DM cups, high quality evidence of their (cost)effectiveness is lacking. This study aims to perform a trial to fill this gap in knowledge. Much of the information needed to judge the effectiveness of DM cups is already incorporated in the Dutch Arthroplasty Register (LROI). This register lends itself perfectly for a nested RCT towards this aim.

**Objective:** The primary objective is to investigate whether there is a difference in the number of hip dislocations following primary total hip arthroplasty (THA), using the posterolateral approach, with a DM cup compared to a unipolar cup in elderly patients 1 year after surgery. The secondary objectives are: to investigate whether there is a difference in the number of revisions; to investigate what the cost-effectiveness and cost-utility is of a DM cup compared to a unipolar cup at 1 year follow-up; to investigate whether there is a difference in the number of hip dislocations and revisions between a DM cup and a unipolar cup 2 years after surgery; to investigate whether there is a difference in patient reported outcomes between a DM cup compared to a unipolar cup 1 and 2 years after surgery; to compare the number of hip dislocations, revisions and PROM data between patients in the randomized DM group and patients in an observational cohort DM group. Finally, long-term survival of DM and unipolar cups will be evaluated based on revision and mortality data registered in the LROI. **Study design:** Prospective multi-center international wide within the European Union (EU), single blinded RCT, nested in the national registry.

**Study population:** Patients ≥ 70 years old, undergoing an elective primary THA. **Intervention (if applicable)**: The intervention group receives a THA with a dual mobility cup, the control group receives a THA with a unipolar cup.

**Main study parameters/endpoints:** Primary: The number of dislocations. Secondary: costs, patient reported outcomes and implant survival.

**Nature and extent of the burden and risks associated with participation, benefit and group relatedness:** In addition to the benefits from regular care, the primary hip arthroplasty procedure, patients might benefit from randomization to receiving a DM cup. DM cups are designed to reduce the risk of hip dislocation, compared to a unipolar cup. Patients may undergo more thorough follow-up than non-study patients and may benefit from this increased surveillance compared with regular care. The only burden associated with study participation is the time needed to complete the cost questionnaires (all other outcomes are part of standard care).

## INTRODUCTION AND RATIONALE

Dislocation is the leading reason for early revision surgery. In the Netherlands, 34.5% of all revisions within the first year after surgery, performed between 2010 and 2015, were due to dislocation.^1^

Most dislocations occur during the first year after surgery, of which approximately half within the first three months after operation.^2–5^ Hip dislocation is a major problem that results in reduced functioning and a deterioration in quality of life^6^, especially in patients with recurrent dislocation who often need revision surgery. In order to establish value based health care for patients with hip osteoarthritis, hospitals within the Santeon network asked patients which outcomes are of value to them. The patients indicated that they feared hip dislocation and that its prevention is paramount to them [personal communication]. In addition to the negative consequences of dislocation for the patients, dislocations also increase healthcare costs.^7;8^ A single dislocation was estimated to add 19% to the hospital costs of an uncomplicated THA, and a revision surgery up to 148%.8 With an average cost for a primary THA of about €10.000 in the Netherlands,^9^ this implies additional costs after dislocation that range from €1.900 to €15.000 per case. To address the problem of dislocation, the dual-mobility (DM) cup was developed in France in the 1970’s.10 The DM cup consists of two articulations between three different components; a metallic acetabular shell, a mobile polyethylene liner and a femoral head. The mobile polyethylene liner articulates both with the acetabular shell and the femoral head. This should provide more stability and biomechanically reduce the risk of dislocation.^11–14^ Dislocation rates reported for the DM-cup range from 0 to 4.6%^12;14–18^ which seems slightly lower than the 0.5 to 6% reported for unipolar cups.^19–24^ Also, the use of DM cups for revision surgery in patients with recurrent dislocation has shown promising results.^2;25;26^ The Dutch national arthroplasty registry (LROI) shows that in 2015 3.9% of all cemented cups was a DM cup.^27^ These DM cups are mainly used in patients with specific characteristics, such as cognitive impairment (not able to follow restrictions after surgery) or neuromuscular diseases (spasms) or as a standard procedure for revision surgeries due to recurrent dislocations. Internationally, DM cups are used more widespread. For instance, in France DM cups are used in an estimated 30% of all THAs.^28^ Potential disadvantages of DM cups are loosening, intra prosthetic dislocation and psoas impingement.^29;30^ In the Netherlands, most DM cups are placed because of specific patient characteristics.^31^ These characteristics might negatively affect the risk for dislocation and revision surgery compared to the general THA population. Nevertheless, several studies reported similar failure rates between DM and unipolar cups.^12;14;17;32^

Beside the type of hip implant used, surgical approach is known to affect the outcome of THA, including dislocations. Different surgical approaches have been developed and each has (potential) advantages and disadvantages. Currently, the posterolateral approach is used in 60% of all THAs in the Netherlands.^33^ Advantages of this approach are good exposure and the preservation of abductor muscles. A disadvantage is that this approach seems to be associated with relatively high dislocation rates compared to other surgical approaches.^21;34–37^ However, with a soft tissue repair this might be diminished.^38^ Other often used approaches are the straight lateral and direct anterior approach. They have a slightly lower risk for dislocation, but have also some disadvantages. The straight lateral approach is associated with abductor insufficiency, resulting in limping.^39;40^ The direct anterior approach has a longer surgical learning curve and a higher risk of complications like nerve injury, periprosthetic fractures and malpositioning of the stem.^41–45^ Considering the disadvantages of other approaches, the posterolateral approach is often preferred. Therefore, many patients could benefit from interventions aimed at reducing dislocation risk after THA using the posterolateral approach.

Despite the increased and, in some countries, broad use of DM cups, high quality evidence of their effectiveness is lacking.^16^ Recent reviews did not identify any randomized controlled trial (RCT) comparing DM cups with unipolar cups.^16;46;47^ The existing studies are of low methodological quality and most of these were performed in France. In France DM cups are used in a broad population. So far only one cost-effectiveness study has been performed, also in France, showing that the DM cup results in cost savings compared with a unipolar cup.^28^ The quality of this study is also limited, largely because it is based on the previous mentioned effectiveness studies of weak quality. Additionally, the results of this cost-effectiveness study are not transferrable outside France.

In conclusion, randomized controlled trials are needed to establish the effectiveness and cost-effectiveness of DM cups for primary THA. This study aims to perform such a trial. As much of the information needed to judge the effectiveness and cost-utility of DM cups is already incorporated in the LROI, this register lends itself perfectly for a nested RCT towards this aim.

The purpose of this study is to investigate whether there is a difference in the number of dislocations after THA, using the posterolateral approach, with a DM cup or a unipolar cup in elderly patients. Moreover, we will perform a cost effectiveness and cost-utility analysis from a health care and societal perspective. Finally, we will compare patient reported outcomes between both groups. As the posterolateral approach is most frequently used but also associated with a (relatively) high dislocation rate, this study will focus on patients who are treated using that surgical approach.

## 1. OBJECTIVES

### Primary objective

To investigate whether there is a difference in the number of hip dislocations following primary total hip arthroplasty (THA), using the posterolateral approach, with a dual-mobility (DM) cup compared to a unipolar cup in elderly patients 1 year after surgery.

### Secondary

- To investigate whether there is a difference in the number of revisions 1 year after surgery.
- To investigate what the cost-effectiveness and cost-utility of a DM cup is, compared to a unipolar cup after primary THA, from the health care and societal perspective at 1 year follow-up.
- To investigate whether there is a difference in the number of hip dislocations and revisions following primary THA with a DM cup compared to a unipolar cup 2 years after surgery.
- To investigate whether there is a difference in patient reported outcomes following primary THA with a DM cup compared to a unipolar cup 1 and 2 years after surgery.
- To compare long-term (5 and 10 year) implant survival based on revision and mortality data from national joint registries.
- To compare the number of hip dislocations, revisions and PROM data between patients in the randomized DM group and patients in an observational cohort DM group.

## 2. STUDY DESIGN

### Design

A prospective multi centre single blinded randomised controlled trial, nested in the national registry of orthopaedic implants to compare the number of hip dislocations following primary total hip arthroplasty (THA), using the posterolateral approach, with a dual-mobility (DM) cup compared to a unipolar cup.

All patients will be followed-up until 2 years after surgery, and after final study follow up, participants remain traceable in the national joint registry for evaluation of long-term survival and mortality.

A third arm (non-randomised) consists of patients that are not eligible for a unipolar cup and therefore will receive a dual mobility cup. See figure 1 for an overview.

**Figure 1.**
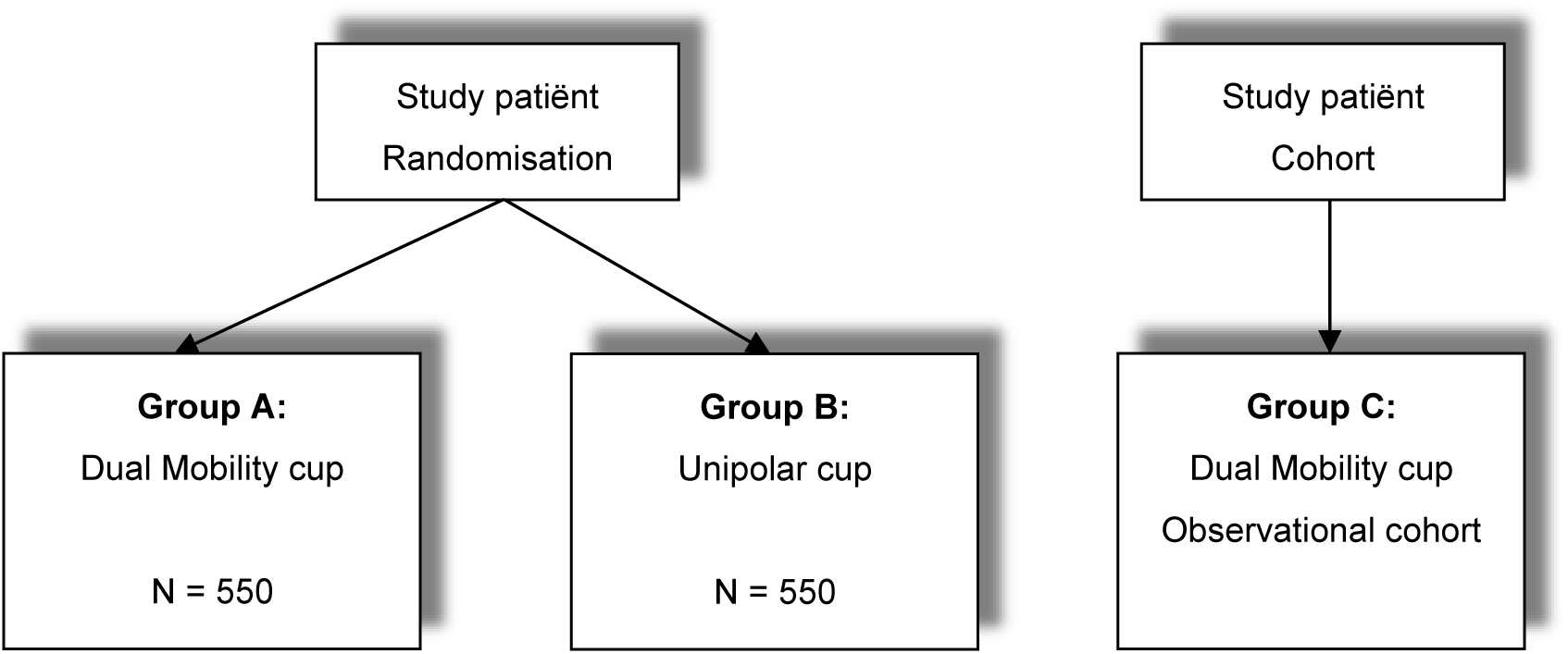
Flowchart with the 3 study groups.

## 3. STUDY POPULATION

### 3.1 Population (base)

All patients at the outpatient clinics of participating centers, that meet the criteria to undergo an elective primary THA will be screened for the in- and exclusion criteria.

### 3.2 Inclusion criteria

To increase the generalizability of the results, wide inclusion criteria are used.

In order to be eligible to participate in this study, a subject must meet all of the following criteria:

Inclusion criteria:

- Patients who are eligible for elective primary THA with a cup, with a 32mm or 36mm liner, for any indication.
- THA using posterolateral surgical approach.
- Patients ≥70 years old
- Adequate comprehension of written and spoken Dutch, Spanish or Swedish

### 3.3 Exclusion criteria

A potential subject who meets any of the following criteria will be excluded from participation in this study:

Exclusion criteria:

- Patients unable to complete PROMs
- Patients with dementia, epilepsy*, spasticity*, mental retardation or alcoholism.

- If dementia or mental retardation is not already mentioned in the medical chart, this can be determined by doctors opinion.
- Patients not eligible for either a unipolar or a DM cup

### 3.4 Sample size calculation

Exact dislocation rates in the Netherlands are unknown. Based on previous studies and reviews, we assume that current dislocation rate for unipolar cups is 4% whereas DM cups result in 1% dislocation.^10;12;14;15;17;18;46^ Power analysis indicates that a total sample of 976 (488 in each group) is needed to detect a difference in dislocations between 4% in the unipolar cup group and 1% in the dual-mobility cup group, using the chi-square test with 80% power and α=0.05. To account for loss to follow-up, 550 patients will be included in each group.

## 4. TREATMENT OF SUBJECTS

Randomized trial:

- <u>Treatment group A:</u> 550 patients will receive a THA with a dual mobility cup.
- <u>Treatment group B:</u> 550 patients will receive a THA with a unipolar cup.

### 4.1 Investigational product/treatment

THA with a dual-mobility cup vs. THA with a unipolar cup.

There are no restrictions to a specific brand of implant. Participating hospitals can use the implants of the industries they usually work with. This study does not investigate any specific product, but rather the concept of DM cups. Both DM and unipolar cups have >95% 5 year survival rates and are commonly used in standard orthopedic care.30

## 5. METHODS

### 5.1 Study parameters/endpoints

#### 5.1.1 Main study parameter/endpoint

The total number of dislocations, regardless of type of treatment (i.e. closed repositioning or revision).

#### 5.1.2 Secondary study parameters/endpoints (if applicable)

- Revision surgery of any component for any reason
- Patient Reported Outcome Measures (PROMs) *The following PROMs are already collected in the LROI according to follow-up of the Dutch orthopaedic association (NOV). This standard follow-up occurs pre- operative, 3 months and 1 year postoperative. For this study one extra measurement will be done at 2 years postoperative*.
  - Physical functioning of the hip, measured by:

- Hip disability and Osteoarthritis Outcome Score Physical Short form (HOOS-PS)48
  - Quality of life, measured by:

- EuroQol 5 Dimensions (EQ-5D)49
  - Pain, measured by:

- Numeric Rating Scale (NRS) for pain in rest rest and during weight bearing
  - Change in functioning, measured by:

- An anchor question about change in functioning.
  *<u>Added as extra question to the standard PROMs:</u>*
  - Fear of hip dislocation on a 5 point Likert scale

- Added at all follow-up moments
  - Healthcare and societal costs related to hip dislocation or surgery.

- Added at 3 months and 1 year postoperative.
  - Education level

- Added as extra question to the PROMs at baseline: “What is the highest education level you achieved?” Answer options according to CBS classification.
  - Awareness of type of cup that was placed

- Added as extra question to the PROMs at all follow-up moments: “Do you know what type of cup was placed?”, “If yes, how did you get this information?”
- Long term (5and 10year) implant survival and mortality based on national registry data.

#### 5.1.3 Other study parameters (if applicable)

Covariates are:

**-** Sex

**-** Age

**-** ASA score

**-** BMI

**-** Brand cup

**-** Type of stem

**-** Type of anesthesia (general or spinal)

**-** Education level

**-** Awareness of type of cup that was placed

### 5.2 Randomisation, blinding and treatment allocation

After signing informed consent, the patients will be randomized in one of the two study groups. A total of 1100 Patients will be randomized into 2 groups: dual-mobility cup versus unipolar cup. Each group consists of 550 patients. The investigator (or his designated representative) will perform the randomization using an online randomization program (CASTOR). Variable randomization blocks of 2, 4 and 6 patients will be used, and we will stratify for center.

Patients will be blinded for group allocation. The principal investigator and the participating surgeons may divert from the randomization scheme based on intra-operative findings.

Any deviation from the assigned treatment group will be reported as a deviation from the protocol.

Patients in treatment group C will take part in the cohort study. These patients are not eligible for a unipolar cup and will receive a dual mobility cup.

### 5.3 Study procedures

During the pre-operative visit at the outpatient clinic, patients who are potential candidates for this study will be screened to determine if they meet the inclusion / exclusion criteria. If the patient is eligible, the investigator (or his designated representative) informs the patient about the study and proposes study participation, according to GCP guidelines. If desired, an additional telephone appointment can be arranged. Patients must sign an ethical committee (METC) approved study informed consent form (ICF) prior to participating in any study specific activities. The ICF can be signed face to face by the patient and investigator, or the patient can sign the ICF remotely and send it to the investigator by mail (due to COVID-19 restrictions).

Subsequently, the investigator can sign the ICF and send one version to the patient. The ICF will be stored in a locked closet in the participating center. Pre-operative data will be collected including: demographics and medical history, NRS for pain in rest and during weight bearing, HOOS-PS and EQ-5D. The patients are also asked to fill out questionnaires. See table 1 for an overview of all measurements and follow-up moments.

**Table 1:**
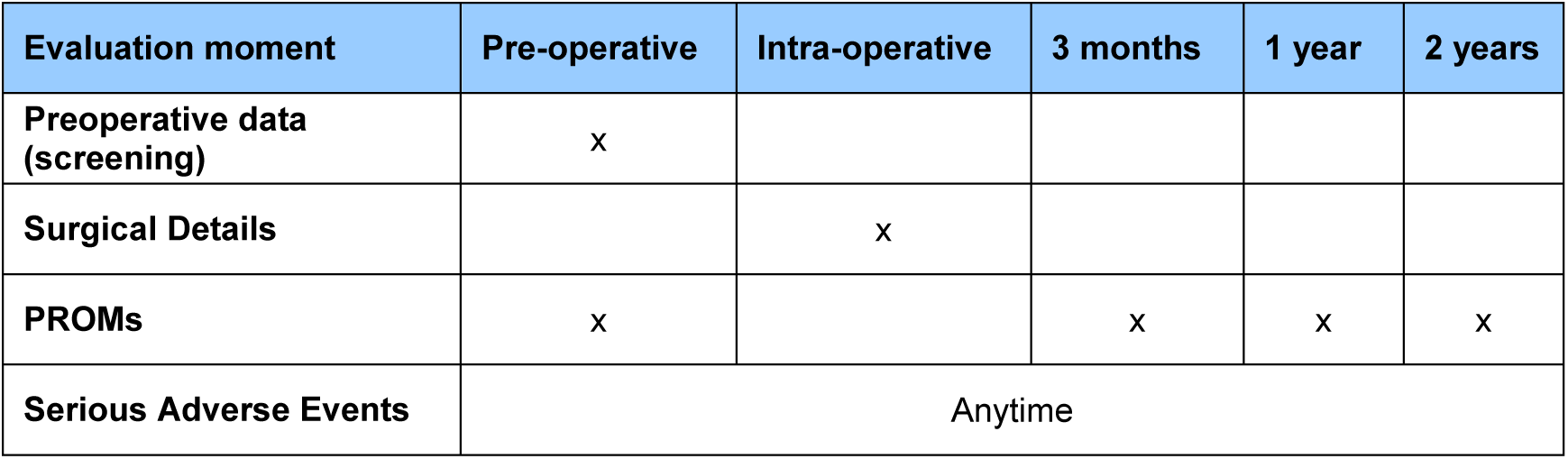
Overview of follow-up moments.

In addition to the above-mentioned questionnaires, patients receive a newsletter that includes a link to a questionnaire that is used to determine the order of importance of the composite outcomes by ranking which composite outcome they consider most relevant. In addition, they perform pairwise comparisons of the different composite outcomes. Completion of the questionnaire is voluntary and all responses are processed anonymously.

### 5.4 Withdrawal of individual subjects

Subjects can leave the study at any time for any reason if they wish to do so, without any consequences. They will be asked for the reason for withdrawal, but do not have to answer if they do not want to. Furthermore, the investigator can decide to withdraw a subject from the study for urgent medical or other reasons.

When a patient withdraws from the study, all data collected prior to the moment of withdrawing will be used for study analysis, unless the patient also withdraws consent for use of this data.

#### 5.4.1 Specific criteria for withdrawal (if applicable)

Not applicable

### 5.5 Replacement of individual subjects after withdrawal

Subjects who withdraw from the study will not be replaced, as long as a minimum of 488 patients per group remains. Otherwise, additional subjects will be recruited.

### 5.6 Follow-up of subjects withdrawn from treatment

The study data of withdrawn patients will be used until the moment of drop-out, unless a patient objects to this.

Patients will be treated according to the best medical judgment of the orthopaedic surgeon, regardless of the study protocol or withdrawal from the study.

### 5.7 Premature termination of the study

Because the devices used in this study are CE marked and will be used according to its labeling, there are no preconceived reasons for premature termination of the study. Upon the sponsor / principal investigator’s decision to terminate or suspend the study, the involved parties and METC will be notified promptly, stating the reasons.

## 6. SAFETY REPORTING

### 6.1 Temporary halt for reasons of subject safety

In accordance to section 10, subsection 4, of the WMO, the sponsor as represented by the principal investigator will suspend the study if there is sufficient ground that continuation of the study will jeopardise subject health or safety. The principal investigator will notify the accredited METC without undue delay of a temporary halt including the reason for such an action. The study will be suspended pending a further positive decision by the accredited METC. The investigator will take care that all subjects are kept informed.

### 6.2 AEs, SAEs and SUSARs

#### 6.2.1 Adverse events (AEs)

Adverse events are defined as any undesirable experience occurring to a subject during the study, whether or not considered related to the implants. Only relevant adverse events reported spontaneously by the subject or observed by the investigator or his staff will be recorded. The following AE’s are directly related to the surgical procedure, and will therefore not be recorded by the investigator: nausea, headache, pain, haemorrhage and wound leakage

#### 6.2.2 Serious adverse events (SAEs)

A serious adverse event is any untoward medical occurrence or effect that

- results in death;
- is life threatening (at the time of the event);
- requires hospitalisation or prolongation of existing inpatients’ hospitalisation;
- results in persistent or significant disability or incapacity;
- is a congenital anomaly or birth defect; or

(S)AEs that are related to a previous known or unknown disease, or conditions that cannot be related to the procedure (like accidents or related to other interventions) will not be recorded. An elective hospital admission will not be considered as a serious adverse event.

The investigator will report the potentially study-related SAEs, as defined above, through the web portal *ToetsingOnline* to the accredited METC that approved the protocol, within 7 days of first knowledge of the investigator for SAEs that result in death or are life threatening followed by a period of maximum of 8 days to complete the initial preliminary report. All other SAEs will be reported within a period of maximum 15 days after the investigator has first knowledge of the serious adverse events.

#### 6.2.3 Suspected unexpected serious adverse reactions (SUSARs)

Not applicable.

### 6.3 Annual safety report

Not applicable.

### 6.4 Follow-up of adverse events

All AEs will be followed until they have abated, or until a stable situation has been reached. Depending on the event, follow up may require additional tests or medical procedures as indicated, and/or referral to the general physician or a medical specialist. SAEs need to be reported till end of study within the Netherlands, as defined in the protocol

### 6.5 [Data Safety Monitoring Board (DSMB) / Safety Committee]

The additional risk of the use of the Dual Mobility cups over and above the risks of standard care, are deemed to be negligible and therefore no DSMB will be established.

## 7. STATISTICAL ANALYSIS

### 7.1 Primary study parameter(s)

The primary outcome, the difference in number of dislocations in both groups, will be analysed using the chi-square test. In case the assumptions of this test are not met, Fischer exact test will be applied. Multilevel logistic regression analysis will be used for analyses in which we adjust for clustering of data (e.g. at the hospital level), possible confounding and effect modification. A multilevel survival model will be used to analyse the survival of the implant, corrected for covariates.

The best way to handle missing values will be defined and applied for all analyses, including the cost-effectiveness analysis.

### 7.2 Secondary study parameter(s)

Secondary study parameters concern revision surgery, and patient reported physical functioning, quality of life, pain, satisfaction, fear of hip dislocation, healthcare and societal costs, device-related complications and reoperations. The secondary outcomes will be analysed using similar multilevel models as appropriate for each outcome (linear/logistic/survival).

### Cost-effectiveness analysis

An economic evaluation will be performed from the societal perspective, for dislocation and Quality Adjusted Life Years (QALYs). Prevailing guidelines of Zorginstituut Nederland will be observed. All costs and consequences relevant to THA, hip dislocation and hip revision will be taken into account.

To compare costs between groups, confidence intervals around the mean differences in costs at one year after THA will estimated using the bias-corrected and accelerated bootstrap method. To account for possible clustering of data and to adjust for possible confounders, multilevel analyses will be performed. To graphically present the incremental cost-effectiveness ratios and uncertainty around them, bootstrapped cost-effect pairs will be plotted on cost-effectiveness planes. Cost-effectiveness acceptability curves will present the probability that the dual-mobility cup is more cost-effective than the unipolar cup for a range of willingness-to-pay thresholds. To study the robustness of these results, sensitivity analyses will be performed.

### 7.3 Other study parameters

#### A priori subgroup analysis

We will analyse patient’s characteristics known for influencing dislocation rate. These include:

- Gender
- Age
- ASA classification
- Femoral head avascular necrosis
- Acute Fracture
- Late posttraumatic condition of the hip

We trust randomization to balance these confounders in both treatment and intervention group.

### 7.4 Interim analysis (if applicable)

Interim analyses for the primary study outcome will be performed when 200 patients have reached the 3 months postoperative PROM evaluation point. In the interim analyses the number of dislocations in each group will be compared. A chi-square test will be used and in case the assumptions of this test are not met, Fischer exact test will be applied. To guard against a type 1 error, we use the O’Brien-Fleming approach. As only one interim analysis will be performed, the alpha for this analysis is set at 0.005. Testing will be done two-sided. Furthermore, we will take the number of revisions and SAE’s in each group in consideration, but will not formally test for differences in these. Results of the interim analysis will be discussed with the steering committee, Van Rens Foundation and the ethical committee. In case of a statically significant and relevant higher number of dislocations in the DM group, or more revisions or SAE’s, appropriate actions will be taken (such as an early termination of the study).

### 7.5 Pre-specified additional win odds analysis

#### 7.5.1 Rationale for a pre-specified additional win odds analysis

Based on advancing methodological insights in clinical trial analysis, we have identified an opportunity to gain a more nuanced and clinically meaningful understanding of the treatment effects by adding a supplementary analysis. The pre-planned chi-square test, while valid, is limited as it treats dislocation as a simple binary event and cannot incorporate the clinical severity of other important outcomes, such as revision surgery or mortality.

The win odds method has emerged as a powerful, patient-centric statistical approach that overcomes these limitations.^50,52,56^ It allows for the evaluation of a hierarchical composite of multiple outcomes, ranked by clinical importance. This provides a more holistic assessment of the net benefit of an intervention. Therefore, the win odds analysis will be performed as an a priori specified additional analysis to enrich the trial’s findings.

#### 7.5.2 Relationship to the Original Primary Analysis

All data required for the win odds analysis are already being collected within the scope of the present protocol. The original primary analysis (chi-square test on dislocation incidence) will be maintained and reported as the formal answer to the original primary research question, and all secondary outcomes will be analysed and reported as described in the present protocol, including the multilevel modelling of secondary clinical endpoints and the full cost-effectiveness analysis.

The win odds analysis will be conducted and reported as a separate, pre-specified additional analysis. This dual approach ensures full adherence to the original protocol while supplementing it with modern, powerful methods to maximize the insights gained from the data generously provided by our trial participants. The analysis is determined prospectively, before completion of follow-up, database lock and any unblinding of treatment-allocated results. This ensures that the analysis is based on methodological merit and not influenced by the observed data, thereby safeguarding scientific integrity.

#### 7.5.3 Order of importance of the composite outcomes

The order of importance of the composite outcomes will be determined in a patient-centered way. Participants included in the REDEP trial receive an online questionnaire at a minimum of 9-months follow-up. A link to the survey will be included in a general study newsletter that will be distributed between December 2025 and February 2026.

Completion of the questionnaire is voluntary and all responses are processed anonymously. In the questionnaire, participants rank which composite outcome they consider most relevant. In addition, they perform pairwise comparisons of the different composite outcomes. The following composite outcomes are included in the questionnaire:

##### Event outcomes

- Revision surgery within 12 months: The revision of any prosthetic component for any reason.

- Dislocation within 12 months: Any dislocation event requiring closed or open reduction.

- Prosthetic Joint Infection within 12 months: Any deep or superficial infection involving the prosthetic joint.

##### Patient Reported Outcome Measures (PROMs)

- Quality of life at 12 months: Measured by the EQ-5D index score.
- Physical functioning at 12 months: Measured by the HOOS-PS total score.

All-cause mortality within 12 months, modelled as time-to-event outcome, will be included as most important in the order of composite endpoints to account for the competing risk of death and prevent immortal time bias. The order of importance for the rest of the composite endpoints is based on the results of the questionnaire, using a structured ranking procedure. The primary determinant will be the cumulative relevance ranking provided by the participants. The pairwise comparisons will be used to validate and refine the hierarchy by assessing the strength and consistency of preferences between outcomes, ensuring that the hierarchy reflects patient-reported relevance and allowing for transparent decision making.

#### 7.5.4 Win odds analysis plan

The additional win odds analysis is based on the win ratio principles outlined by Pocock et al.^50^ Brunner, Vandemeulebroecke & Mütze modified these principles in their win odds analysis to incorporate ties, thereby reducing the risk of overestimating the treatment effect.^51^

The win odds analysis compares all possible pairs of patients between the DM cup group and the UP cup group, e.g. every patient in the DM cup group is compared to every patient in the UP cup group. Within each pair, we evaluate composite outcomes in descending order of importance until one patient of the pair shows a superior outcome compared to the other patient. If the comparison of the most important and therefore first composite outcome is inconclusive, then the second-most important composite outcome is compared between the two patients in the pair. If the patient in the DM cup group has the better outcome it is called a ‘win’. If the patient in the UP cup group has the better outcome it is called a ‘loss’. If the comparison of all composite outcomes is inconclusive it is called a ‘tie’. The win odds is then determined by the following formula:

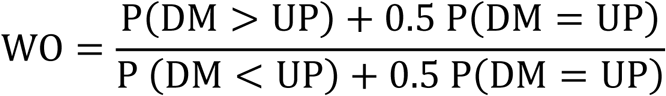

P (X > Y) = the percentage of wins
P (X < Y) = the percentage of losses
P (X = Y) = the percentage of ties

A win odds larger than 1 favors the DM cup group and a win odds smaller than 1 favors the UP cup group. The 95% confidence interval and p-value will be calculated using the WinRatio package in R57, which is based on the large-sample inference methods described by Bebu & Lachin.^55^

For the event outcomes the time-to-event determines a ‘win’ or ‘loss’ in each pair. For example, if the patient in the DM cup group suffered a dislocation at 90-days postoperatively and the patient in the UP cup group suffered a dislocation at 120-days postoperatively, it is considered a ‘win’.

For the PROMs a ‘win’ or ‘loss’ is determined by the largest within-patient improvement. For example, if the HOOS-PS total score of the patient in the DM cup group improved by 20 points and the HOOS-PS total score of the patient in the UP cup group improved by 25 points, it is considered a ‘loss’. If the comparisons of all composite outcomes are inconclusive, it is considered a ‘tie’.

#### 7.5.5 Clinically meaningful margins in determining wins and losses

Some win odds study examples suggest determining a ‘win’ or ‘loss’ only if the difference between the patients in the pair exceeds a clinically meaningful quantity, or margin.^52, 56^ However, there are statistical arguments against the use of such margins as they increase the number of ‘ties’ in the analysis, thereby reducing statistical power. Additionally, imposing a margin is inconsistent with the principle of rank-based statistical testing. On the other hand, there is a clinical rationale for the use of such margins, since a single point difference in a PROM or a single day difference in time-to-event is not clinically meaningful. Consequently, our primary win odds analysis will not include any margins, but we will perform a supplementary analysis from a clinical perspective including clinically meaningful margins. For this supplementary analysis, we will include the following margins:

For the event outcomes a margin of 30 days will be applied. For any event outcome, it is considered a ‘win’ if a patient in the DM cup groups remains event-free for at least 30 days longer than the patient in the UP cup group within the 12-month follow-up.

For the PROMs the margins are based on the minimal important change values derived from the literature. For the HOOS-PS total score, a margin of 9.47 is considered clinically meaningful, as described by Emara et al.53 This indicates that it is considered a ‘win’ if the within-patient improvement in HOOS-PS total score of the patient in the DM cup group is at least 9.47 points larger than the within-patient improvement of the patient in the UP cup group. For the EQ5D index score, a margin of 0.071 is considered clinically meaningful, as described by Silva et al.54. This indicates that it is considered a ‘win’ if the within-patient improvement in EQ5D index score of the patient in the DM cup group is at least 0.071 points larger than the within-patient improvement of the patient in the UP cup group.

## 8. ETHICAL CONSIDERATIONS

### 8.1 Regulation statement

This study will be conducted according to the principles of the Declaration of Helsinki (2013) and in accordance with the Medical Research Involving Human Subjects Act (WMO) and Good Clinical Practice guidelines.

### 8.2 Recruitment and consent

During the pre-operative visit, patients that are possible candidates for this study will be screened to determine if they meet the inclusion / exclusion criteria. If the patient is eligible, the investigator (or his designated representative) will propose participation in the study to the patient, according to GCP guidelines.

Patients must sign an METC approved study informed consent form prior to participating in any study related activities. The patients will be given adequate time to consider their decision (>1 week).

### 8.3 Objection by minors or incapacitated subjects (if applicable)

*Not applicable*.

### 8.4 Benefits and risks assessment, group relatedness

In addition to the benefits from the primary hip arthroplasty procedure e.g. reduced pain, improved range of motion, patients might benefit from the type of acetabular cup that is used in the intervention group. Patients in the intervention group receive a dual mobility acetabular cup. Dual mobility cups are designed to reduce the risk of hip dislocation, compared to a unipolar cup.

### 8.5 Compensation for injury

The sponsor/investigator has a liability insurance which is in accordance with article 7 of the WMO. The implants (dual mobility cup and unipolar cup) used in this study are concepts of standard care, widely used in the Netherlands. Therefore we will apply for exemption for the insurance for subjects participating in this study.

The sponsor also has an insurance which is in accordance with the legal requirements in the Netherlands (Article 7 WMO). This insurance provides cover for damage to research subjects through injury or death caused by the study.

The insurance applies to the damage that becomes apparent during the study or within 4 years after the end of the study.

### 8.6 Incentives (if applicable)

Patients will not receive any (financial) compensation for participation in this study.

## 9. ADMINISTRATIVE ASPECTS, MONITORING AND PUBLICATION

### 9.1 Handling and storage of data and documents

Data will be handled confidentially and anonymously. Each subject will be given an identification code and only research personnel involved in the logistics of the study will have access to the subject identification code list which can be used to link the data to the subject. The code is based on consecutive numbers. The handling of personal data will comply with the Dutch Personal Data Protection Act (in Dutch: Algemene Verordening Gegevensbescherming, AVG). Data will be kept for 15 years after the end of the study. This includes digital information of the study parameters and digital PROMs. Digital information will be kept in Castor EDC. PROMs sent by mail are kept on paper with only a study number.

### 9.2 Monitoring and Quality Assurance

Monitoring of the study will take place during the total study duration, in the investigating centre, according to guidelines set by the initiating center (OLVG).

### 9.3 Amendments

Amendments are changes made to the research after a favourable opinion by the accredited METC has been given. All amendments will be notified to the METC that gave a favourable opinion.

All substantial amendments will be notified to the METC and to the competent authority. Non-substantial amendments will not be notified to the accredited METC and the competent authority, but will be recorded and filed by the sponsor.

### 9.4 Annual progress report

The sponsor/investigator will submit a summary of the progress of the trial to the accredited METC once a year. Information will be provided on the date of inclusion of the first subject, number of subjects included and number of subjects who have completed the trial, serious adverse events/ serious adverse reactions, other problems, and amendments.

### 9.5 Temporary halt and (prematurely) end of study report

The investigator/sponsor will notify the accredited METC of the end of the study within a period of 8 weeks. The end of the study is defined as the last patient’s last questionnaire. The data collection in the national joint registry will continue, even after a temporary halt or (prematurely) end of this study.

The sponsor will notify the METC immediately of a temporary halt of the study, including the reason of such an action.

In case the study is ended prematurely, the sponsor will notify the accredited METC within 15 days, including the reasons for the premature termination.

Within one year after the end of the study, the investigator will submit a final study report with the results of the study, including any publications/abstracts of the study, to the accredited METC.

### 9.6 Public disclosure and publication policy

All publications and other public disclosures of the research data by the investigators will be made independent from the subsidizing party. The subsidizing party will be informed about publication at least one month before submitting a publication.

## 10. STRUCTURED RISK ANALYSIS

There is minimal risk associated with participating in this study over and above that of the primary hip arthroplasty procedure. Serious complications may be associated with any total joint replacement surgery. These complications include, but are not limited to: infection; genitourinary disorders; gastrointestinal disorders; vascular disorders, including thrombus; bronchopulmonary disorders, including emboli; myocardial infarction or death. All devices are CE marked and will be used according to its labelling. Patients will be treated in the best medical judgment of the surgeon, regardless of the study protocol. Assessment involves questionnaires and anterior-posterior and lateral radiographs. The patient’s burden from the study consists of extra questions added to questionnaires at the standard LROI follow-up moments (5 minutes extra at 3 months and 1 year) and one extra questionnaire at 2 year follow-up (15 minutes extra). In addition to the benefits from the primary hip arthroplasty procedure e.g. reduced pain, improved range of motion, patients might benefit from the type of acetabular cup that is used in this study. Patients in the intervention group receive a dual mobility cup, instead of a unipolar acetabular cup. Dual mobility cups are designed to reduce the risk of hip dislocation.

## LIST OF ABBREVIATIONS AND RELEVANT DEFINITIONS

AE: Adverse Event
CV: Curriculum Vitae
DM: Dual Mobility
DSMB: Data Safety Monitoring Board
EQ-5D: EuroQol 5 Dimension
GCP: Good Clinical Practice
HOOS-PS: Hip disability and Osteoarthritis Outcome Score Physical Short form
IB: Investigator’s Brochure
IC: Informed Consent
LROI: Dutch Arthroplasty Register (in Dutch: Landelijke Registratie Orthopedische Implantaten)
METC: Medical research ethics committee (MREC); in Dutch: medisch ethische toetsing commissie (METC)
RCT: Randomized Controlled Trial
(S)AE: (Serious) Adverse Event
Sponsor: The sponsor is the party that commissions the organisation or performance of the research, for example a pharmaceutical company, academic hospital, scientific organisation or investigator. A party that provides funding for a study but does not commission it is not regarded as the sponsor, but referred to as a subsidising party
SUSAR: Suspected Unexpected Serious Adverse Reaction
THA: Total Hip Arthroplasty
Wbp: Personal Data Protection Act (in Dutch: Wet Bescherming Persoonsgevens)
WMO: Medical Research Involving Human Subjects Act (in Dutch: Wet Medisch-wetenschappelijk Onderzoek met Mensen

## Data Availability

All data produced in the present work are contained in the manuscript

## Notes

### Competing Interest Statement

The authors have declared no competing interest.

### Clinical Trial

NCT04031820

### Summary of Updates

The author list was adjusted to include all authors that contributed to this version of the study protocol.

### Funding

- Landelijk Register Orthopedische Interventies (formerly: Van Rens Fonds)

